# Cutaneous tactile sensation and standing balance in children with autism: A Preliminary Report

**DOI:** 10.1101/2022.09.05.22279614

**Authors:** Komal Kukkar, Pranav J. Parikh, Chyung Fen-Kao, Sambit Mohapatra

**Affiliations:** Center for Neuromotor and Biomechanics Research, Department of Health and Human Performance, University of Houston, Houston, Texas; Cuomo First Step Early Childhood Center, Heartshare Human Services of New York, New York, NY; The Department of Rehabilitation and Movement Science, The University of Vermont, Burlington, Vermont

**Keywords:** ASD, tactile, balance, posture

## Abstract

**BACKGROUND:** Autistic Spectrum Disorder (ASD) presents with a multitude of problems such as physical, social, emotional, psychological, etc. Most common physical problems are impairments in standing balance and posture. It is unknown whether these impairments have any association between tactile sensation or are purely due to deficits in sensory processing and integration. We hypothesized that foot tactile sensation in ASD is positively correlated to performance in standing balance as measured by Pediatric Balance scale.

**METHODS:** The data collected at Heartshare Human Services of New York was used for secondary analysis. It consisted of 12 participants and included: 1. Muscle and joint ROM testing to rule out any muscle involvement in balance problems. 2. Tactile sensation testing at four sites on sole of foot bilaterally using Semmes Weinstein monofilament. 3. Pediatric Balance Scale (PBS) for balance testing.

**RESULTS:** We found significant positive correlation between cutaneous tactile sensation (SWF) and Pediatric Balance scale (PBS) measures in our participants i.e., reduced tactile sensation was moderately associated with impaired balance score.

**CONCLUSIONS:** We propose that during conventional clinical assessment for individuals with ASD, foot tactile sensation should not be overlooked, and included as a part of somato-sensory assessment. In addition, enhancing foot tactile sensation could also be used for targeted interventions to improve balance in children with ASD.

## 1.0 INTRODUCTION

Autism spectrum disorders (ASDs) is a life-long developmental disorder of the brain affecting nearly 2 in 1000 children in the United States (Bhat AN1, Landa RJ, 2011; Rapin I1, 1998). The average lifetime cost associated with direct medical expenses, rehabilitation costs, long-term care costs and special education amounts to nearly USD 3.2 million per person (Alcantara et al., 2011). Although early diagnosis of children with ASD is critical, it is often delayed until school age (Mandell et al., 2005). ASD can be classified into three types: a). Autistic disorder or classic autism; b). Asperger’s syndrome, a mild form of autistic disorder; and c). Pervasive developmental disorder or atypical autism, which meets some of the criteria for autistic disorder or asperger syndrome. Neuroanatomic, pathologic, and neurobehavioral studies have suggested abnormalities in premotor (Mostofsky & Ewen, 2011), cerebellar, and parietal cortices (Haas et al., 1996) and in the thalamocortical connectivity in children with ASD leading to sensorimotor, sociocommunicative, and cognitive impairments (Nair A1, Treiber JM, Shukla DK, Shih P, 2013).

Children with ASD have a range of difficulties affecting the initiation, organization, and performance of movements leading to a range of movement problems (Maski et al., 2011). Most children with ASD present with problems maintaining balance while standing or during locomotion (Graham SA1, Abbott AE, Nair A, Lincoln AJ, Müller RA, n.d.; Stins & Emck, 2018). Maintenance of an upright stance requires continuous processing of visual, somatosensory, and vestibular systems to assess the stability of the body, and initiation of anticipatory and/or corrective neuro-muscular adjustments when challenged to keep the body’s center of mass within the base of support (Horak, 2006; Shumway-Cook & Woollacott, 2000). Children with ASD when compared to age-matched typically developing children have been found to demonstrate an impaired ability to maintain an upright stance during static and dynamic tasks (Fournier KA1, Hass CJ, Naik SK, Lodha N, 2010; Mosconi, Wang, et al., 2015). These autistic children tend to rely more on sensory information for the control of balance. That is, when sensory information is blocked or removed, children with ASD demonstrate increased postural sway (Kohen-Raz et al., 1992; Minshew et al., 2004). Importantly, balance deficits in these children are more prominent during dynamic challenging tasks that require increased reliance on sensory information than a static task (Wang et al., 2016; Wang Z1, Hallac RR2, Conroy KC3, White SP3, Kane AA2, Collinsworth AL2, Sweeney JA4, 2016). Impaired balance control in autistic children affects mobility and independence in activities of daily living, and may affect the ability to acquire manipulatory skills (Noterdaeme et al., 2002).Therefore, it is important to understand the underlying mechanisms.

Poor ability to control the balance may result from sensory processing deficits as well as impaired sensorimotor integration in ASD (Lim YH1, Partridge K2, Girdler S2, 2017). Difficulty processing somatosensory information including touch is common in ASD (Caminha & Lampreia, 2012). These children may show little or no response to sensory stimulation (Randell et al., 2019) or show heightened response to touch (Espenhahn et al., 2021). In addition, they have an impaired ability to localize or discriminate tactile stimulation problems (He et al., 2021; Hense et al., 2019), especially when in presence of another sensory stimulation such as visual distractors (Poole, 2018). Despite significant deficits in processing tactile information, there is a considerable lack of research understanding the contribution of impaired tactile sensation especially in the foot to balance deficits in ASD. In this secondary data analysis, we hypothesized that reduced tactile sensation in the foot will be associated with poor balance control as suggested by lower score on the Pediatric Berg Balance (PBS) scale in individuals with ASD.

## 2.0 MATERIALS AND METHODS

### 2.1 Data

This is a secondary data which was obtained from Heartshare Human Services of New York preschool. This data was collected at Heartshare by a physical therapist under the supervision of the head of the department as a part of standard routine of care. In this preliminary report, we included participants between 4 to 6 years of age with a diagnosis of autism spectrum disorder (ASD) and a history of imbalance or falls. Exclusion criteria included tightness in calf muscle and hip flexors or knee hyperextension. Twelve participants were met inclusion/exclusion criteria. This study was deemed exempt from Institutional Review Board (IRB) review by the IRB at the University of Vermont/University of Vermont Health Network (Exempt Criteria 45 CFR 46.104(d)(4)(ii)). The information was utilized by the authors in such a manner that the identity of the participants could not readily be ascertained directly or through identifiers linked to the participants, the authors did not contact the participants, and the authors will not re-identify participants.

### 2.2 Data Collection

According to the department head, the following procedure was followed to collect the data:

#### Assessment of Muscle tightness and Range of Motion

##### Calf muscles

The participants were instructed to lie down prone on a therapy table with hands on the side. The PT placed the knee in 90 degrees of flexion and the ankle in a neutral position. Goniometer was placed on the ankle joint with fulcrum on the lateral malleolus with the proximal arm aligned to the lateral midline of fibula with fibular head as reference and the distal arm aligned to the lateral aspect of 5^th^ metatarsal. The ankle was passively moved into dorsiflexion along with the distal arm of goniometer to measure dorsi-flexion range of motion. After this, the knee was extended, and the procedure was repeated to measure the ankle dorsi-flexion range of motion.

##### Hip flexors

Participants were instructed to lie down in the supine position on a therapy table with both knees on the edge of the table. The Thomas test was performed to assess tightness in the hip flexors. One knee was moved towards the chest and the participant was instructed to hold that position using both hands and let the other leg go down. This procedure was repeated with the opposite knee. If the leg that was not held by the subject reaches parallel to the therapy table or goes beyond, it ruled out hip flexor tightness of that leg.

##### Knee extensors

Participants were instructed to lie supine with both legs on a therapy table. Goniometer fulcrum was placed on the lateral epicondyle of femur with proximal arm along the lateral aspect of femur using greater trochanter for reference and the distal arm along the lateral aspect of fibula with the fibular head and lateral malleolus as reference. Then, the PT pushed the knee into hyperextension along with goniometer’s distal arm with a hand placed under the ankle to measure the knee hyperextension range of motion.

#### Assessment of Cutaneous Tactile Sensation

This involved testing for cutaneous tactile sensation at four sites - great toe (GT), first metatarsal head (MT1), fifth metatarsal head (MT5) and heel (H) - on the plantar surface of each foot using a Semmes–Weinstein 5.07 (10g) monofilament. The inability to sense 5.07 SWM is indicative of the loss of protective sensation (Ünver & Akbaş, 2018). Participants laid flat with their back on a mat. Both feet were cleaned with cleaning wipes and labeled as 1(GT), 2(MT1), 3(MT5) and 4(H). A couple of practice trials were performed to ensure the participants understood the testing process. Participants were instructed to keep their eyes closed before the filament touched the foot. The filament was touched with sufficient pressure applied to bend it for one second with the participants lying down. After filament was touched, participants were instructed to look at foot and either point out or number the site where the filament was touched. The examiner performed 24 trials (three trials at each of the four sites on either feet) in a random order. At each site, one trial was a “sham” in which the examiner did not touch the participant, but still asked for response. One point was awarded for each correct response (including a “no” response for a sham trial) for a total cutaneous tactile score of up to 24 points (12 points per foot).

#### Functional/Clinical Outcome Measures

Balance was assessed with the Pediatric Balance Scale (PBS), which is a 14-item performance-based assessment of balance related tasks (Casey et al., 2015). The PBS scale is a common clinical tool to assess balance and mobility and has been shown to have a strong positive correlation with the dynamic balance and gait ability (Evkaya et al., 2020). Each task is scored on an ordinal scale from 0 (unable) to 4 (independent). The sum of all scores was used as the final outcome score from a total score of 56.

### 2.3 Data Analysis

Data obtained was analyzed using SPSS software, version 24 (IBM, Inc, Chicago, IL). Descriptive data is presented in Table 1. Range of motion measurements were reported separately for either limb. Comparisons of cutaneous tactile sensation (SWF) scores were made with measures of balance (PBS) by Pearson’s correlation test. We classified the results according to the correlation coefficient (r) - very strong (r>0.9), strong (r from 0.7 to 0.9), moderate (r from 0.5 to 0.7), and weak (r from 0.3 to 0.5). Statistical significance was calculated at p<0.05.

**Table 1.** Participant information.

| Patients | Tactile Sensibility | Balance Score | Knee Extension |  | Ankle Dorsiflexion with Knee Extension |  | Ankle Dorsiflexion with Knee Flexion |  |
| --- | --- | --- | --- | --- | --- | --- | --- | --- |
|  |  |  | Left | Right | Left | Right | Left | Right |
| 1 | 24.00 | 50.00 | -3.00 | -2.00 | 10.00 | 10.00 | 20.00 | 20.00 |
| 2 | 19.00 | 48.00 | -2.00 | -2.00 | 15.00 | 15.00 | 39.00 | 38.00 |
| 3 | 14.00 | 50.00 | -2.00 | -1.00 | 8.00 | 7.00 | 25.00 | 25.00 |
| 4 | 21.00 | 48.00 | 0.00 | 0.00 | 10.00 | 10.00 | 25.00 | 25.00 |
| 5 | 22.00 | 49.00 | 3.00 | 5.00 | 8.00 | 10.00 | 10.00 | 15.00 |
| 6 | 16.00 | 50.00 | 12.00 | 12.00 | 6.00 | 6.00 | 12.00 | 12.00 |
| 7 | 11.00 | 51.00 | 0.00 | 0.00 | 5.00 | 5.00 | 12.00 | 12.00 |
| 8 | 14.00 | 40.00 | 0.00 | 0.00 | 6.00 | 3.00 | 10.00 | 10.00 |
| 9 | 8.00 | 34.00 | 0.00 | 0.00 | 8.00 | 8.00 | 16.00 | 16.00 |
| 10 | 10.00 | 40.00 | -2.00 | -2.00 | 10.00 | 8.00 | 20.00 | 20.00 |
| 11 | 11.00 | 44.00 | 2.00 | 5.00 | 10.00 | 10.00 | 20.00 | 18.00 |
| 12 | 22.00 | 47.00 | 10.00 | 13.00 | 6.00 | 6.00 | 20.00 | 22.00 |

## 3.0 RESULTS

### 3.1 Goniometry: Range of Motion

Ankle ROM: With knee in 90 degrees of flexion, the average ROM for the left ankle was 19.08 ± 8.229 (mean ± SD) degrees and the average ROM for the right ankle was 19.42 ± 7.669 degrees. With knee fully extended, the average ROM for the left ankle was 8.5 ± 2.75 degrees and the average ROM for the right ankle was 8.17 ± 3.12 degrees (**Table 1**).

Knee extension: Knee extension were tested bilaterally to rule out any knee hyperextension. The average ROM for the left knee hyperextension was 1.5 ± 4.78 degrees and the average ROM for the right knee hyperextension was 2.33 ± 5.31 degrees (**Table 1**).

### 3.2 Cutaneous Tactile Perception (SWF test)

The average (± SD) cutaneous tactile sensation score on SWF test was 16.00 ± 5.461 out of a total possible score of 24 (**Table 1**).

### 3.3 Functional measures

*Pediatric Berg Balance Scale (PBS)*

The average PBS score was 45.92 ± 5.316 out of a total possible score of 56 (**Table 1**).

### 3.4 Correlations

We found significant moderate correlation (r=0.604) between cutaneous tactile sensation (SWF) and Pediatric Berg Balance scale (PBS) score in our participants with ASD (r=0.604; p=0.037). Thus, reduced tactile sensation demonstrated moderate association with impaired balance (**Fig. 1**) explaining approximately 37% of the variance.

**Figure 1.**
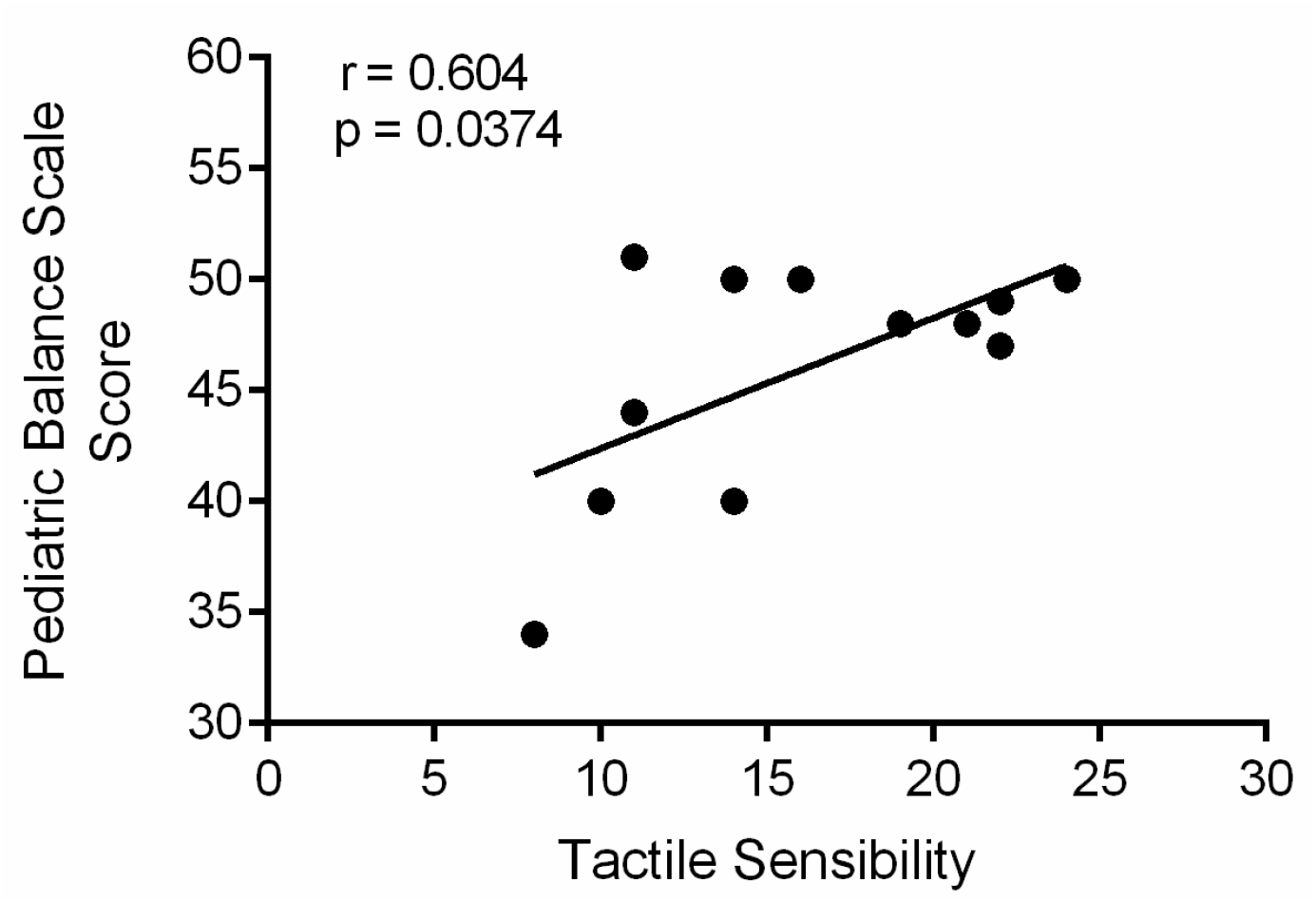
Correlation between foot tactile sensation and the score on the Pediatric Berg Balance (PBS) scale. The tactile sensation measure explained inter-individual differences in the PBS score (r=0.604; p=0.037).

## 4.0 Discussion

Balance control is impaired in children with ASD. These impairments have been suggested to be due to sensorimotor abnormalities, which are also common in ASD and among the earliest manifestations of the disorder (Mosconi, Mohanty, et al., 2015). Individuals with ASD have severe difficulties in the integration of perceived stimuli into a meaningful entity (van Berckelaer-Onnes, 2004). Our secondary data analysis examined the relationship between cutaneous tactile sensation and standing balance. We found a moderate positive correlation between foot tactile sensation as measured by SWF and the clinically relevant balance function from the score on PBS.

### 4.1 Cutaneous tactile sensation in ASD

Impaired plantar cutaneous tactile sensation contributes to deficits in standing balance. This has been shown in individuals with neurological diseases such as stroke(Parsons et al., 2016) and multiple sclerosis (Citaker et al., 2011). In ASD, hypersensitivity or hyposensitivity to tactile sensation may result from inability to filter irrelevant sensory information. Whereas some studies have shown alteration of tactile stimuli in both adults and children with autism (Blakemore et al., 2006), others have shown tactile detection to be normal in autism (Cascio et al., 2008). These differences were primarily due to the differences in type and location of tactile stimulations. The data obtained consisted of the locations which were standardized for sensory testing in all the participants. It included four specific sites: Great Toe-GT, 1^st,^ and 5^th^ Metatarsals-MT-1, MT-5, and Heel-H. Integrity of sensation in these regions is particularly relevant to maintenance of standing balance and posture. Also, a standardized clinical tool was used to assess tactile sensation, which is tested at a specific load (i.e., 10g) that discriminates intactness of protective tactile sensation. This was important as we were only looking at the association between these sensations and standing balance, which is also contributed by various other sensations.

### 4.2 Cutaneous Tactile sensation and its relationship with balance

We found the tactile sensation explained 36% of the variation in the clinical balance score. This finding is consistent with a study done by Cruz et al on elderly where they found that foot tactile sensation strongly correlates with BBS (Cruz-Almeida Y1, Black ML2, Christou EA3, 2014), as well as with study by Pamela et al concluding strong correlation between SWME and lower limb physical performance (Carrer et al., 2018). This is important because poor balance potentially leads to frequent falls and secondary complications. Consequently, enhancing tactile sensation could become a targeted intervention to improve balance, and thus reduce falls. Thus, tactile sensation should not be overlooked and should be added to clinical assessment protocols for balance.

One limitation of this analysis was small sample size. A larger study is needed to confirm our observation. Also, results of our analysis should be interpreted carefully as this is a correlation between sensation and balance, but does not suggest causation.

## Data Availability

All data produced in the present study are available upon reasonable request to the authors

## 5.0 Implications and Future Directions

Relationship between foot tactile sensation on feet and standing balance in individuals with ASD is crucial. This secondary analysis examined cutaneous tactile sensation in individuals with ASD and found that tactile sensation is most reduced in ASD individuals who have difficulties maintaining standing balance. The results of this analysis would be helpful for refining clinical assessment protocols so that somatosensory screening can be added to motor assessment protocols. In addition, ASD is a sensory processing disorder and future research should attempt at differentiating tactile sensation deficits with sensory processing problems so that an approximate causation can be reached, and targeted interventions can be delivered.

## Conflict of Interest

All authors declare no conflict of interests, and were fully involved in the secondary data analysis and preparation of the manuscript. The material within has not been and will not be submitted for publication.

## Notes

Declarations of interest: None.

### Competing Interest Statement

The authors have declared no competing interest.

### Funding Statement

This study did not receive any funding

### Author Declarations

This study was deemed exempt from Institutional Review Board (IRB) review by the IRB at the University of Vermont/University of Vermont Health Network (Exempt Criteria 45 CFR 46.104(d)(4)(ii)). The information was utilized by the authors in such a manner that the identity of the participants could not readily be ascertained directly or through identifiers linked to the participants, the authors did not contact the participants, and the authors will not re-identify participants.

## Literature Cited

Alcantara, J., Alcantara, J. D., & Alcantara, J. (2011). A systematic review of the literature on the chiropractic care of patients with autism spectrum disorder. Explore: The Journal of Science and Healing, 7(6), 384–390. https://doi.org/10.1016/j.explore.2011.08.001

Bhat AN1, Landa RJ, G. JC. (2011). Current perspectives on motor functioning in infants, children, and adults with autism spectrum disorders.

Blakemore, S. J., Tavassoli, T., Calò, S., Thomas, R. M., Catmur, C., Frith, U., & Haggard, P. (2006). Tactile sensitivity in Asperger syndrome. Brain and Cognition, 61(1), 5–13. https://doi.org/10.1016/j.bandc.2005.12.013

Caminha, R. C., & Lampreia, C. (2012). Findings on sensory deficits in autism: Implications for understanding the disorder. Psychology and Neuroscience, 5(2), 231–237. https://doi.org/10.3922/j.psns.2012.2.14

Carrer, P., Trevisan, C., Curreri, C., Giantin, V., Maggi, S., Crepaldi, G., Manzato, E., & Sergi, G. (2018). Semmes-Weinstein Monofilament Examination for Predicting Physical Performance and the Risk of Falls in Older People: Results of the Pro.V.A. Longitudinal Study. Archives of Physical Medicine and Rehabilitation, 99(1), 137-143.e1. https://doi.org/10.1016/j.apmr.2017.08.480

Cascio, C., McGlone, F., Folger, S., Tannan, V., Baranek, G., Pelphrey, K. A., & Essick, G. (2008). Tactile perception in adults with autism: A multidimensional psychophysical study. Journal of Autism and Developmental Disorders, 38(1), 127–137. https://doi.org/10.1007/s10803-007-0370-8

Casey, A. F., Quenneville-Himbeault, G., Normore, A., Davis, H., & Martell, S. G. (2015). A Therapeutic Skating Intervention for Children with Autism Spectrum Disorder. Pediatric Physical Therapy, 27(2), 170–177. https://doi.org/10.1097/PEP.0000000000000139

Citaker, S., Gunduz, A. G., Guclu, M. B., Nazliel, B., Irkec, C., & Kaya, D. (2011). Relationship between foot sensation and standing balance in patients with multiple sclerosis. Gait & Posture, 34(2), 275–278. https://doi.org/10.1016/j.gaitpost.2011.05.015

Cruz-Almeida Y1, Black ML2, Christou EA3, C. D. (2014). Site-specific differences in the association between plantar tactile perception and mobility function in older adults.

Espenhahn, S., Godfrey, K. J., Kaur, S., Ross, M., Nath, N., Dmitrieva, O., McMorris, C., Cortese, F., Wright, C., Murias, K., Dewey, D., Protzner, A. B., McCrimmon, A., Bray, S., & Harris, A. D. (2021). Tactile cortical responses and association with tactile reactivity in young children on the autism spectrum. Molecular Autism, 12(1). https://doi.org/10.1186/s13229-021-00435-9

Evkaya, A., Karadag-Saygi, E., Karali Bingul, D., & Giray, E. (2020). Validity and reliability of the Dynamic Gait Index in children with hemiplegic cerebral palsy. Gait and Posture, 75, 28–33. https://doi.org/10.1016/j.gaitpost.2019.09.024

Fournier KA1, Hass CJ, Naik SK, Lodha N, C. J. (2010). Motor coordination in autism spectrum disorders: a synthesis and meta-analysis.

Graham SA1, Abbott AE, Nair A, Lincoln AJ, Müller RA, G. DJ. (n.d.). The Influence of Task Difficulty and Participant Age on Balance Control in ASD.Graham SA1, Abbott AE, Nair A, Lincoln AJ, Müller RA, G. D. (n.d.). The Influence of Task Difficulty and Participant Age on Balance Control in ASD.

Haas, R. H., Townsend, J., Courchesne, E., Lincoln, A. J., Schreibman, L., & Yeung-Courchesne, R. (1996). Neurologic abnormalities in infantile autism. Journal of Child Neurology, 11(2), 84–92. https://doi.org/10.1177/088307389601100204

He, J. L., Wodka, E., Tommerdahl, M., Edden, R. A. E., Mikkelsen, M., Mostofsky, S. H., & Puts, N. A. J. (2021). Disorder-specific alterations of tactile sensitivity in neurodevelopmental disorders. Communications Biology, 4(1). https://doi.org/10.1038/s42003-020-01592-y

Hense, M., Badde, S., Köhne, S., Dziobek, I., & Röder, B. (2019). Visual and Proprioceptive Influences on Tactile Spatial Processing in Adults with Autism Spectrum Disorders. Autism Research, 12(12), 1745–1757. https://doi.org/10.1002/aur.2202

Horak, F. B. (2006). Postural orientation and equilibrium: What do we need to know about neural control of balance to prevent falls? Age and Ageing, 35(SUPPL.2). https://doi.org/10.1093/ageing/afl077

Kohen-Raz, R., Volkmar, F. R., & Cohen, D. J. (1992). Postural Control in Children with Autism 1. In Journal of Autism and Developmental Disorders (Vol. 22, Issue 3).

Lim YH1, Partridge K2, Girdler S2, M. S. (2017). Standing Postural Control in Individuals with Autism Spectrum Disorder: Systematic Review and Meta-analysis.

Mandell, D. S., Novak, M. M., & Zubritsky, C. D. (2005). Factors Associated With Age of Diagnosis Among Children With Autism Spectrum Disorders. Pediatrics, 116(6), 1480–1486. https://doi.org/10.1542/peds.2005-0185

Maski, K. P., Jeste, S. S., & Spence, S. J. (2011). Common neurological co-morbidities in autism spectrum disorders. In Current Opinion in Pediatrics (Vol. 23, Issue 6, pp. 609–615). https://doi.org/10.1097/MOP.0b013e32834c9282

Minshew, N. J., Sung, K., Jones, B. L., & Furman, J. M. (2004). Underdevelopment of the postural control system in autism.

Mosconi, M. W., Mohanty, S., Greene, R. K., Cook, E. H., Vaillancourt, D. E., & Sweeney, J. A. (2015). Feedforward and Feedback Motor Control Abnormalities Implicate Cerebellar Dysfunctions in Autism Spectrum Disorder. Journal of Neuroscience, 35(5), 2015–2025. https://doi.org/10.1523/JNEUROSCI.2731-14.2015

Mosconi, M. W., Wang, Z., Schmitt, L. M., Tsai, P., & Sweeney, J. A. (2015). The role of cerebellar circuitry alterations in the pathophysiology of autism spectrum disorders. In Frontiers in Neuroscience (Vol. 9, Issue SEP). Frontiers Media S.A. https://doi.org/10.3389/fnins.2015.00296

Mostofsky, S. H., & Ewen, J. B. (2011). Altered connectivity and action model formation in autism is autism. In Neuroscientist (Vol. 17, Issue 4, pp. 437–448). https://doi.org/10.1177/1073858410392381

Nair A1, Treiber JM, Shukla DK, Shih P, M. RA. (2013). Impaired thalamocortical connectivity in autism spectrum disorder: a study of functional and anatomical connectivity.

Noterdaeme, M., Mildenberger, K., Minow, F., & Amorosa, H. (2002). Evaluation of neuromotor deficits in children with autism and children with a specific speech and language disorder. European Child and Adolescent Psychiatry, 11(5), 219–225. https://doi.org/10.1007/s00787-002-0285-z

Parsons, S. L., Mansfield, A., Inness, E. L., & Patterson, K. K. (2016). The relationship of plantar cutaneous sensation and standing balance post-stroke. Topics in Stroke Rehabilitation, 23(5), 326–332. https://doi.org/10.1080/10749357.2016.1162396

Poole, D. (2018). Supplemental Material for Visual-Tactile Selective Attention in Autism Spectrum Condition: An Increased Influence of Visual Distractors. Journal of Experimental Psychology: General. https://doi.org/10.1037/xge0000425.supp

Randell, E., McNamara, R., Delport, S., Busse, M., Hastings, R. P., Gillespie, D., Williams-Thomas, R., Brookes-Howell, L., Romeo, R., Boadu, J., Ahuja, A. S., McKigney, A. M., Knapp, M., Smith, K., Thornton, J., & Warren, G. (2019). Sensory integration therapy versus usual care for sensory processing difficulties in autism spectrum disorder in children: Study protocol for a pragmatic randomised controlled trial. Trials, 20(1). https://doi.org/10.1186/s13063-019-3205-y

Rapin I1, K. R. (1998). Neurobiology of autism.

Shumway-Cook, A., & Woollacott, M. (2000). Attentional Demands and Postural Control: The Effect of Sensory Context. In Journal o/Cerontolngt: MEDICAL SCIENCES (Vol. 55, Issue 1). https://academic.oup.com/biomedgerontology/article/55/1/M10/545748

Stins, J. F., & Emck, C. (2018). Balance performance in autism: A brief overview. In Frontiers in Psychology (Vol. 9, Issue JUN). Frontiers Media S.A. https://doi.org/10.3389/fpsyg.2018.00901

Ünver, B., & Akbaş, E. (2018). Effects of plantar sensitivity on balance and mobility in community-dwelling older adults: A Turkish study. Australasian Journal on Ageing, 37(4), 288–292. https://doi.org/10.1111/ajag.12558

van Berckelaer-Onnes, I. a. (2004). Sixty years of autism. Nederlands Tijdschrift Voor Geneeskunde, 148(21), 1024–1030.

Wang, Z., Hallac, R. R., Conroy, K. C., White, S. P., Kane, A. A., Collinsworth, A. L., Sweeney, J. A., & Mosconi, M. W. (2016). Postural orientation and equilibrium processes associated with increased postural sway in autism spectrum disorder (ASD). Journal of Neurodevelopmental Disorders, 8(1). https://doi.org/10.1186/s11689-016-9178-1

Wang Z1, Hallac RR2, Conroy KC3, White SP3, Kane AA2, Collinsworth AL2, Sweeney JA4, M. M. (2016). Postural orientation and equilibrium processes associated with increased postural sway in autism spectrum disorder (ASD).

